# An Inflammatory Signature Associated with Genetic Predisposition to Acute Necrotizing Encephalopathy

**DOI:** 10.64898/2026.04.24.26350762

**Authors:** Sophie Desgraupes, Stéphanie Boireau, Mirna Khalil, Safa Aouinti, Sébastien Nisole, Karine Bolloré, Wiem Barbaria, Federica Barzaghi, Robertino Dilena, Maartje Boon, Roelineke J. Lunsing, Edouard Tuaillon, Mia Westerholm-Ormio, Kumaran Deiva, Dewi P. Bakker, Taco W. Kuijpers, E. Ann Yeh, Ming Lim, Marie-Christine Picot, Pierre Meyer, Nathalie J. Arhel

## Abstract

**Background:** Acute necrotizing encephalopathy (ANE) is a rare and severe neurologic complication of viral infection in children, thought to result from a hyperacute cytokine storm causing blood-brain barrier disruption and central nervous system injury. Despite characteristic clinical and radiologic features, ANE remains poorly understood at the molecular level, with no validated biomarkers or targeted therapies. We aimed to determine whether genetic predisposition to ANE due to *RANBP2* variants is associated with a distinct immunologic signature.

**Methods:** We conducted a prospective biological study of familial ANE (ANE1, NCT06731790). We included 23 heterozygous carriers of the *RANBP2* c.1754C>T (p.Thr585Met) variant from 10 families, and 28 noncarriers (median age, 40 years [range, 4-72]). Soluble immune mediators, transcriptomic analyses, multiparameter flow cytometry, and cellular imaging were analysed in peripheral blood mononuclear cells (PBMCs) and monocytes. Baseline and resiquimod-stimulated immune responses were analysed within the same statistical model, with genetic status as the primary predictor.

**Findings:** The RANBP2 Thr585Met mutation was associated with a dysregulated inflammatory phenotype characterized by reduced basal mediator production and exaggerated TNF-α responses following stimulation (estimated difference, +2,098 pg/mL; 95% CI, 1,121 to 3,076; P=0.0001). Transcriptomic and flow cytometry analyses showed broad reprogramming of myeloid cells with enrichment of CXCR3-high CD14-high subsets. Expansion of these populations was associated with increased long-term disease burden. The *RANBP2* variant was the only independent factor associated this inflammatory phenotype.

Interpretation: RANBP2-associated ANE is characterised by a distinct immunological signature that can inform disease stratification and support the development of targeted immunotherapeutic approaches.

## Introduction

Surveillance data from the U.S. Centers for Disease Control and Prevention indicate that, over the past two seasons, an unprecedented proportion of pediatric influenza-associated deaths (13%) were accompanied by severe neurologic injury, including acute necrotizing encephalopathy (ANE) and encephalitis^1–3^.

Despite characteristic clinical and radiologic features and emerging diagnostic frameworks^4^, the biological basis of ANE remains poorly defined, with no molecular diagnosis, no validated biomarkers or targeted therapies, resulting in persistent uncertainty regarding prognosis, recurrence risk, and treatment response.

Several case studies have reported elevated inflammatory cytokines in serum or cerebrospinal fluid^5–7^, and early recognition and treatment with anti-inflammatory therapies, such as corticosteroids, intravenous immunoglobulins, and plasmapheresis, have been associated with improved clinical outcomes^5,8,9^. These observations suggest that dysregulated cytokine responses may contribute to ANE pathogenesis, although the underlying mechanism remains unknown.

Although ANE can be sporadic, recurrent disease has been linked to a single point mutation (c.1754C>T; p.Thr585Met) in *RANBP2*, which encodes a nucleoporin component of the nuclear pore complex involved in regulating nucleocytoplasmic transport^10^. To date, however, no studies have examined how this mutation contributes to disease recurrence.

To address this gap, we conducted a prospective clinical study of familial ANE (NCT06731790), profiling peripheral blood mononuclear cells (PBMCs) from 10 affected families, including carriers and noncarriers of the c.1754C>T (p.Thr585Met) mutation, and unrelated healthy controls using multiplex cytokine assays, transcriptomics, multiparameter flow cytometry, and cellular imaging.

## Methods

### Study design and objectives

This exploratory, controlled, pathophysiological pilot study evaluated the inflammatory phenotype associated with heterozygous mutations in *RANBP2* (ANE1) in patients. Given prior evidence that RANBP2 regulates inflammatory signaling and cytokine expression^11^, the study was designed to assess whether *RANBP2* mutations are associated with exaggerated basal and stimulus-induced inflammatory responses.

The primary objective was to compare inflammatory profiles of circulating immune cells from individuals with ANE1 and age- and sex-matched non-carrier controls. Secondary objectives included characterization of *RANBP2* allelic expression. The study was approved by the Comité de protection des personnes Île-de-France VII (approval no. 24.00903.000298).

### Participants and samples

Participants with confirmed *RANBP2* mutations and matched controls were recruited during an international ANE1 meeting^12^. Each participant underwent a single peripheral blood draw performed outside routine clinical care and outside of an acute relapse period for affected individuals, with no subsequent follow-up. Biological samples included whole blood, serum, peripheral blood mononuclear cells (PBMCs), and monocyte-derived macrophages and microglia.

Given the extreme rarity of ANE1, no formal sample size calculation or randomization was performed. Adults and children carrying the *RANBP2* c.1880C>T (p.Thr585Met) mutation, as well as noncarrier controls, were eligible for inclusion. Written informed consent was obtained from all adult participants or from parents or legal guardians for minors. Individuals were excluded if consent could not be obtained, if they were legally incapacitated or deprived of liberty, or if they were participating in another study with an ongoing exclusion period.

Study participants carrying a pathogenic *RANBP2* variant are referred to as mutation carriers (MUT) and are further classified as symptomatic having one or more episode of ANE as reported by primary treating physician and validated by the study team (P.M. and M.L.).

Participants without the *RANBP2* variant are referred to as noncarriers (NC); this group includes non-carrier relatives and unrelated controls.

### Statistical analyses for the primary analysis

Statistical analyses were performed using R software (version 4.5.0). All tests were two-sided, with a prespecified significance level of 0.05. No adjustment was made for multiple comparisons given the exploratory nature of the study.

Descriptive statistics were reported using means and standard deviations or medians and interquartile ranges, depending on data distribution. Categorical variables were summarized as frequencies and percentages.

Individuals with *RANBP2* mutations were matched to noncarrier controls by sex and age epochs (4-13, 14-23, 34-43, 44-53, and 54-74 years). This matching was incorporated into all primary analyses using mixed-effects models with pair as a random effect.

For the primary analysis, differences in inflammatory marker expression between *RANBP2*-mutated participants and matched controls were assessed using linear mixed-effects models, with group (mutated vs. control) as a fixed effect and the matching pair identifier as a random effect. This approach accounted for age- and sex-matching and within-pair correlation. Analyses were conducted separately for basal and stimulated conditions.

### ANE disease impact score

All participants were clinically appraised by an experienced clinician (P.M.). A composite disease impact score (range: 0-14) was devised (M.L. and P.M.) to assess mutation penetrance and disease burden. The score incorporated the number of ANE episodes, cognitive impairment, functional impairment, and long-term support needs, all ascertained at the time of inclusion. Cognitive impairment was assessed qualitatively: 1 (mild), defined by any learning difficulties; 2 (moderate), defined by additional difficulty sustaining or holding a conversation; and 3 (severe), absent or minimal expressive speech. Functional impairment was assessed using the modified Rankin Scale (mRS; range, 0 to 5). Long-term support needs were scored as 1 point for requiring assistance and 2 points for requiring a protected or supported living environment. Scores of zero for functional disability, cognitive severity, or long-term support requirements represent confirmed absence of impairment, not missing data.

For analysis, participants were pragmatically classified into four ordinal severity categories: no disease burden (noncarrier participants), minimal impact (score 0, n=11; carriers without ANE episodes at inclusion), moderate impact (scores 1-3, n=9), or high impact (scores ≥5, n=3). Categories were analyzed as an ordinal variable without assuming equal distances between levels.

## Results

### Participant characteristics

Ten families affected by ANE1 were included in the study (**Figure 1A**). A total of 51 participants were enrolled, including 23 carriers of the *RANBP2* c.1754C>T (p.Thr585Met) mutation, and 28 noncarriers, comprising 12 noncarrier family members and 16 unrelated controls matched for age and sex. Both carriers and noncarrier relatives were represented across all 10 families (**Table 1**).

**Figure 1.**
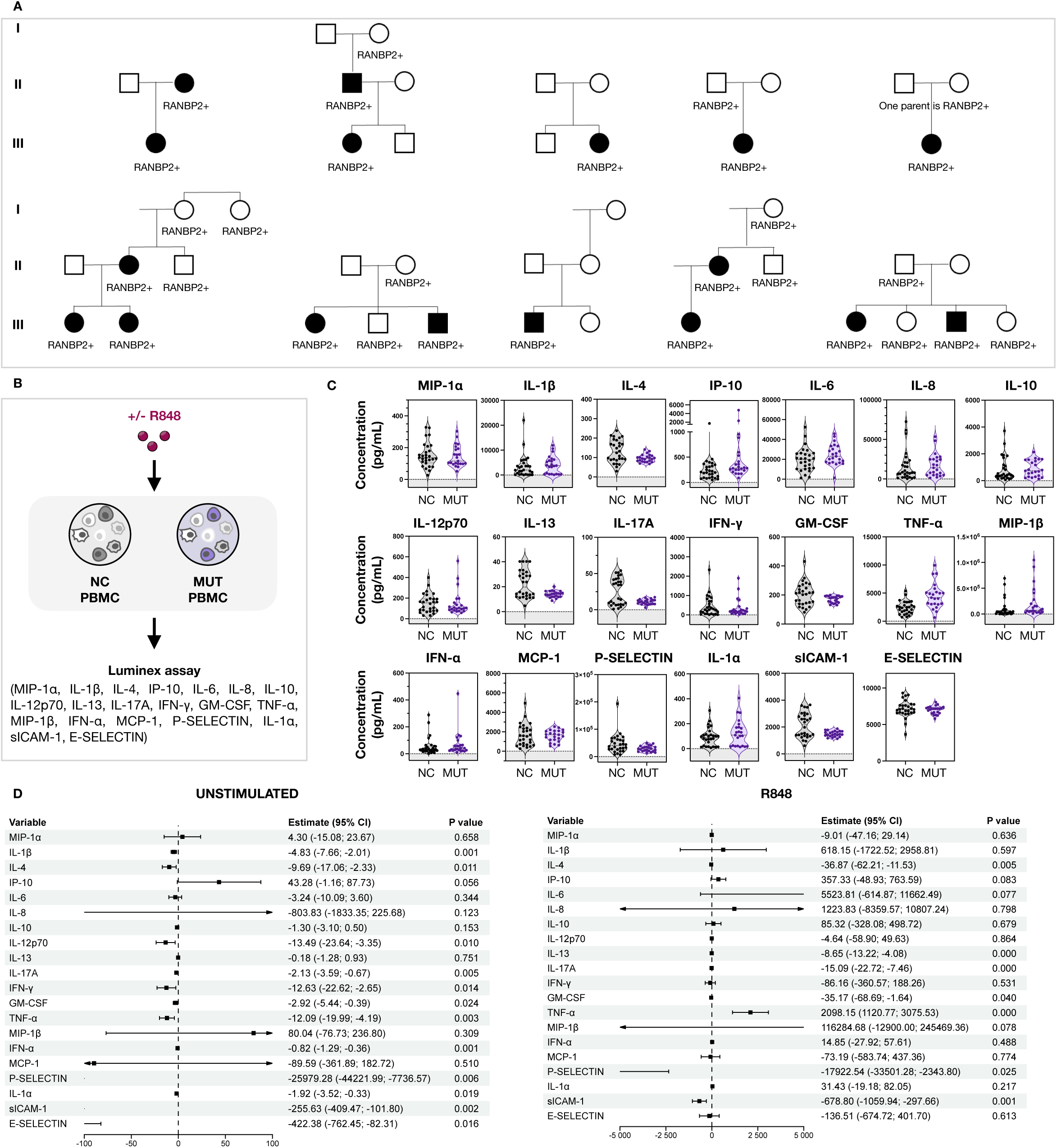
Cytokine secretion by PBMCs from individuals with *RANBP2* mutation at baseline and after innate immune stimulation, as compared with controls. (A) Pedigrees of families with RANBP2-associated ANE. Squares indicate males and circles females. Filled symbols denote individuals with a history of ANE; open symbols denote unaffected individuals. *RANBP2* mutation carriers are indicated by “RANBP2+”; absence of this label indicates noncarriers. All carriers carry the same heterozygous RANBP2 variant (c.1754C>T). In one family, parental carrier status is not shown at the family’s request. (B) Experimental design. Peripheral blood mononuclear cells (PBMCs) from individuals carrying the RANBP2 c.1754C>T (p.Thr585Met) variant (MUT) and age- and sex-matched noncarrier controls (NC) were cultured under unstimulated conditions or stimulated with the TLR7/8 agonist resiquimod (R848). Secreted cytokines and chemokines were quantified by multiplex bead-based immunoassays (Luminex). (C) Violin plots show the distribution of per-participant cytokine concentrations following R848 stimulation. Each point represents an individual participant. Dashed lines indicate medians, and dotted lines interquartile ranges; gray shading denotes the lower limit of assay detection. (D) Statistical analysis. Participants were matched by age group and sex. Group comparisons were performed using linear mixed-effects models with group as a fixed effect and matched pairs as random effects (two-sided α=0.05). Fold values (R848/unstimulated) for all measured mediators are provided in **appendix Figure S2**.

**Table 1.**
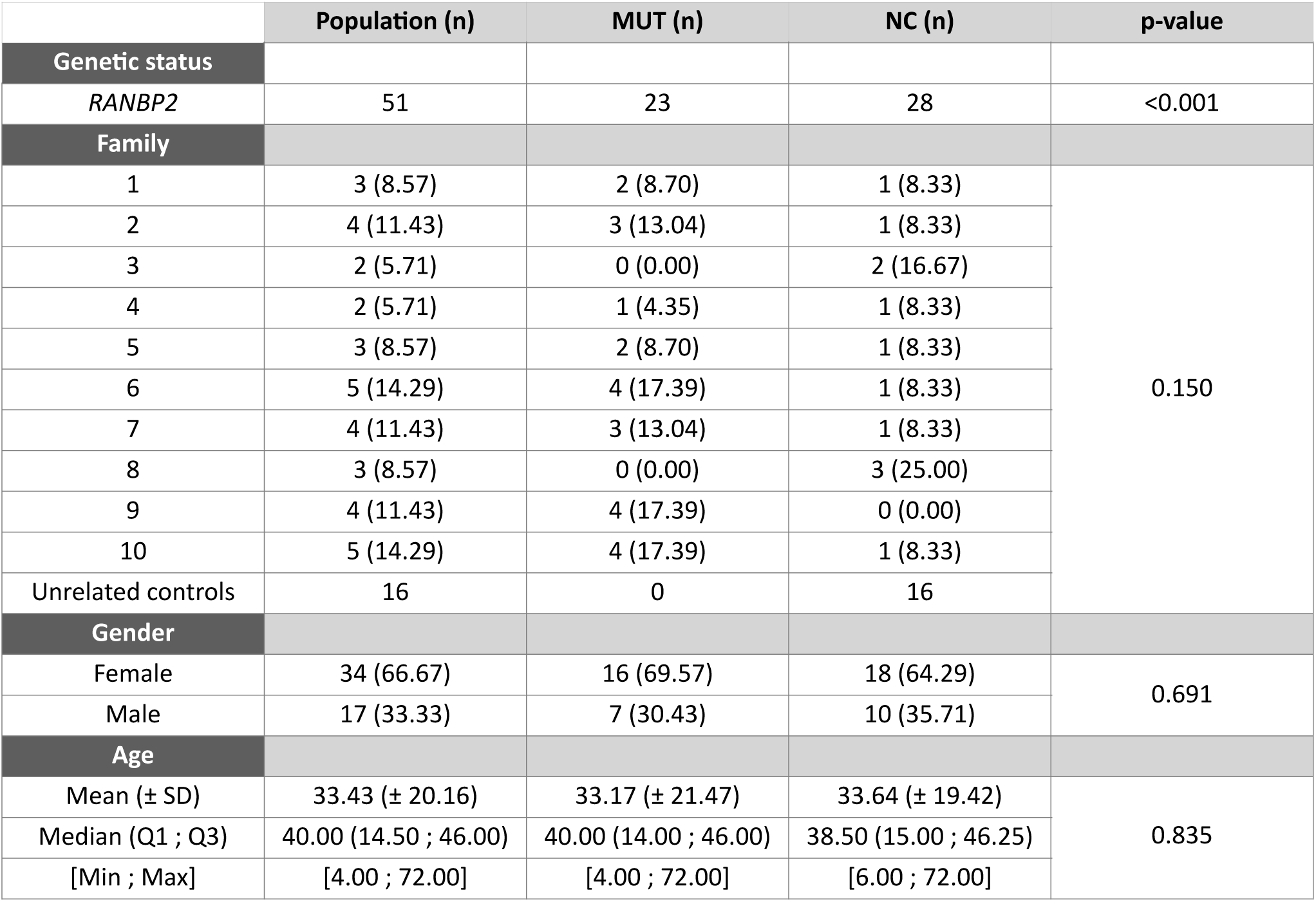
Participant characteristics. Demographic, genetic, and clinical characteristics of study participants, including carriers of the *RANBP2* c.1754C>T (p.Thr585Met) mutation (MUT) and noncarriers (NC), comprising noncarrier family relatives and unrelated controls.

|  | Population (n) | MUT (n) | NC (n) | p-value |
| --- | --- | --- | --- | --- |
| Genetic status |  |  |  |  |
| RANBP2 | 51 | 23 | 28 | <0.001 |
| Family |  |  |  |  |
| 1 | 3 (8.57) | 2 (8.70) | 1 (8.33) | 0.150 |
| 2 | 4 (11.43) | 3 (13.04) | 1 (8.33) |  |
| 3 | 2 (5.71) | 0 (0.00) | 2 (16.67) |  |
| 4 | 2 (5.71) | 1 (4.35) | 1 (8.33) |  |
| 5 | 3 (8.57) | 2 (8.70) | 1 (8.33) |  |
| 6 | 5 (14.29) | 4 (17.39) | 1 (8.33) |  |
| 7 | 4 (11.43) | 3 (13.04) | 1 (8.33) |  |
| 8 | 3 (8.57) | 0 (0.00) | 3 (25.00) |  |
| 9 | 4 (11.43) | 4 (17.39) | 0 (0.00) |  |
| 10 | 5 (14.29) | 4 (17.39) | 1 (8.33) |  |
| Unrelated controls | 16 | 0 | 16 |  |
| Gender |  |  |  |  |
| Female | 34 (66.67) | 16 (69.57) | 18 (64.29) | 0.691 |
| Male | 17 (33.33) | 7 (30.43) | 10 (35.71) |  |
| Age |  |  |  |  |
| Mean (± SD) | 33.43 (± 20.16) | 33.17 (± 21.47) | 33.64 (± 19.42) | 0.835 |
| Median (Q1 ; Q3) | 40.00 (14.50 ; 46.00) | 40.00 (14.00 ; 46.00) | 38.50 (15.00 ; 46.25) |  |
| [Min ; Max] | [4.00 ; 72.00] | [4.00 ; 72.00] | [6.00 ; 72.00] |  |

All mutation carriers and their relatives had received a prior genetic diagnosis, which was an inclusion criterion for the study. Mutation status and heterozygous expression was confirmed by allelic quantitative PCR, as previously reported (**appendix p2**)^13^. All unrelated control participants were confirmed to carry wild-type *RANBP2* (**appendix Figure S1**). One participant had been described in a previous case report^14^.

Among the 23 carriers, 11 (47.8%) had not experienced an ANE episode at enrollment, whereas 9 (39.1%), 1 (4.4%), and 2 (8.7%) had experienced one, two, or three ANE episodes, respectively. Two participants were initially misdiagnosed with Lyme disease and myositis and were later correctly diagnosed with ANE following subsequent episodes. The median age at first, second, and third ANE episodes was 5.5 years (range, 0.25-31), 5.0 years (range, 4.0-10.0), and 10.0 years (range, 5.0-15.0), respectively.

Influenza virus infection was the most commonly identified trigger, reported in 9 of 11 episodes (81.8 %) for which an infectious cause was investigated. Other reported pathogens included bocavirus and parainfluenza virus type 4^14^.

Of the 12 carriers who had experienced one or more ANE episodes, 7 (58.3%) had cognitive and/or motor sequelae at enrollment, whereas 5 (41.7%) had neither. Cognitive involvement (n=6) included slowed cognitive processing, attention deficits, visual disturbances, and anxiety symptoms. Motor deficits (n= 5) were associated with mRS scores of 1 (n=2), 2 (n=2), and 4 (n=1). Six participants (50%) reported functional impairments requiring educational or workplace assistance.

### Increased TNF-α secretion upon stimulation in ANE1 PBMCs

To assess the impact of the RANBP2 Thr585Met mutation on inflammatory responses, PBMCs from mutation carriers and noncarrier controls were analyzed under steady-state conditions and following stimulation with the Toll-like receptor (TLR) 7/8 agonist resiquimod (R848) (**Figure 1B; appendix p2**).

Following innate immune stimulation, PBMCs from mutation carriers secreted significantly higher levels of tumor necrosis factor α (TNF-α) than controls (estimated difference, 2098.1 pg/mL; 95% confidence interval CI, 1120.8-3075.5; P<0.001), whereas no concomitant increases (either statistically unsignificant or too low in magnitude to be considered biologically meaningful) were observed across a broad panel of other inflammatory and T-cell-associated cytokines (**Figure 1C**).

In contrast, under unstimulated conditions, PBMCs from mutation carriers exhibited reduced basal production of TNF-α, IL-1α, and IL-1β compared with controls (**Figure 1D**). The combination of diminished steady-state cytokine production and exaggerated inducible TNF-α secretion resulted in markedly increased relative inflammatory reactivity in mutation carriers (fold difference, 396.3; 95% CI, 251.8 - 540.7; p<0.001) (**appendix Figure S2**).

Together, these findings indicate that the RANBP2 Thr585Met mutation is associated with a dysregulated inflammatory phenotype characterized by reduced basal cytokine production and selective amplification of TNF-α responses following innate immune stimulation, without generalized activation of adaptive immune cytokines.

### Broad transcriptional dysregulation of immune pathways and myeloid compartment skewing in ANE1

To better understand the basis of the altered inflammatory responses associated with the RANBP2 Thr585Met mutation, we performed bulk RNA sequencing. Of 18,736 expressed genes, only 260 genes (1.4%) were differentially expressed between mutation carriers and related noncarriers under unstimulated conditions, and 215 (1.1%) following R848 stimulation, indicating a high degree of transcriptional similarity between the two groups. In both conditions, approximately two-thirds of differentially expressed genes (167 genes under unstimulated condition and 171 genes after stimulation) were downregulated in mutation carriers compared with controls (**Figure 2A, appendix Figure S3A-B; appendix pp 2-3**).

**Figure 2.**
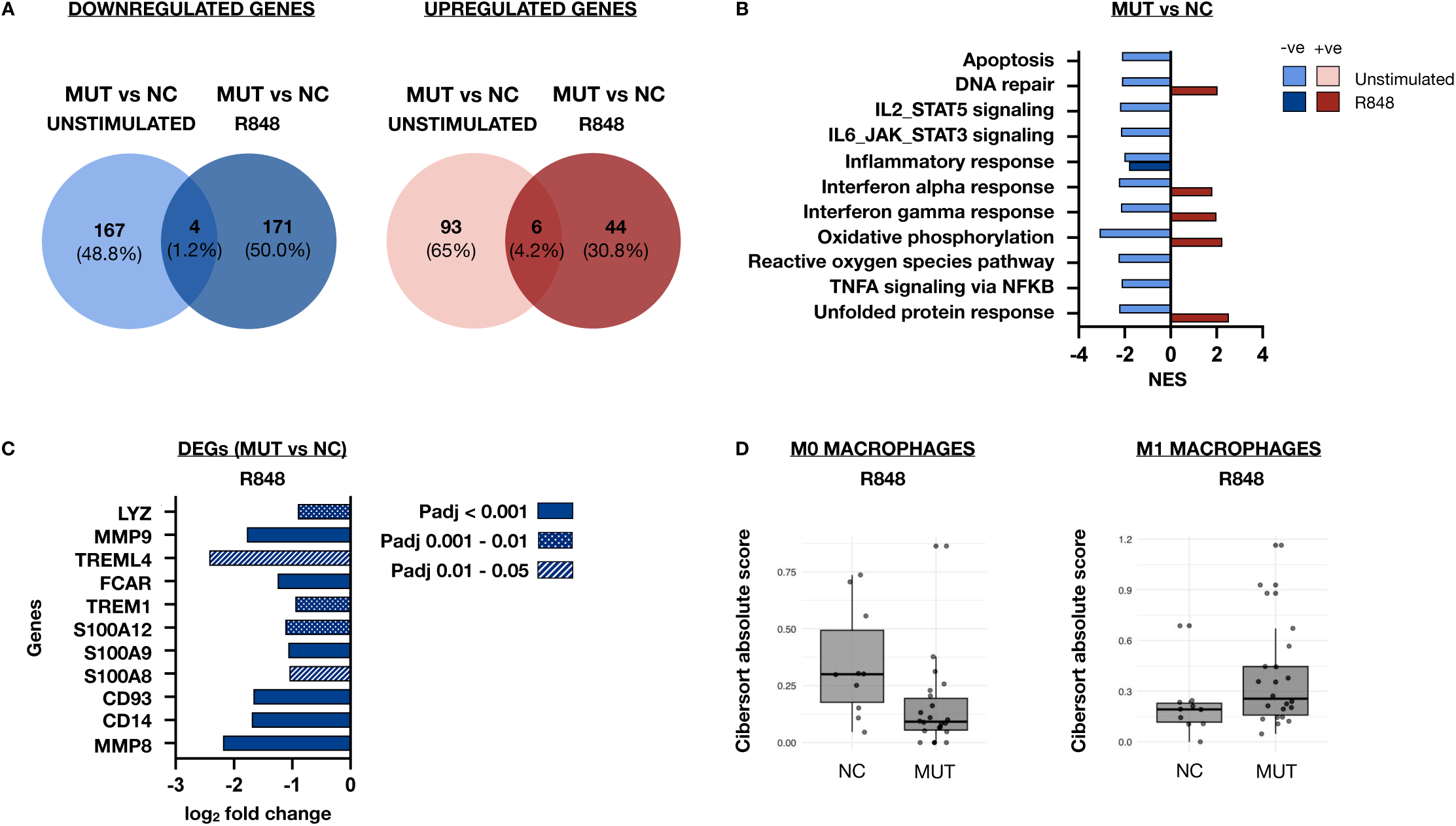
Whole-cell RNA sequencing of PBMCs from individuals with *RANBP2* mutation at baseline and after innate immune stimulation, as compared with controls. (A) Venn diagrams showing differentially expressed genes (DEGs) in *RANBP2* mutation carriers compared with related noncarriers under unstimulated and R848-stimulated conditions. (B) Gene-set enrichment analysis of Hallmark pathways showing positive (+ve) and negative (-ve) enrichment in mutation carriers relative to noncarriers under both conditions; normalized enrichment scores (NES) are shown. (C) Differential expression of genes associated with a classical monocyte transcriptional signature in mutation carriers. (D) Cell-type deconvolution of bulk RNA-sequencing data using CIBERSORTx. RNA-sequencing analyses were performed in mutation carriers and related noncarriers only. Differential expression was assessed using normalized gene counts and linear modeling with adjustment for sex, age, and batch effects. Nominal P values were corrected for multiple testing using the Benjamini-Hochberg method, and significance was defined by a false discovery rate of less than 0.05.

Gene-set enrichment analysis (GSEA) using Hallmark gene sets demonstrated coordinated negative enrichment of immune, inflammatory, and stress-response pathways in unstimulated samples from *RANBP2* mutation carriers compared with wild-type controls. Suppressed pathways included interferon-α and interferon-γ responses, TNF-α-NFκB signaling, IL-2-STAT5 and IL-6-JAK-STAT3 signaling, and inflammatory and stress responses, indicating a broadly attenuated baseline immune and stress-response state, consistent with the reduced cytokine production observed in unstimulated samples (**Figure 2B**).

Following stimulation with R848, this transcriptional profile was inverted, with positive enrichment of interferon-α and interferon-γ responses, and stress-response pathways in *RANBP2* mutation carriers. Among the most prominent changes observed in the R848-stimulated samples, markers of classical monocyte (*MMP8*, *CD14*, *CD93*, *S100A8, S100A9, S100A12*, *TREM1* and *FCAR*) and classically activated monocytes (*TREML4*, *MMP9* and *LYZ*)^15–17^, showed reduced expression in mutation carriers relative to noncarriers (**Figure 2C**).

These data suggest that altered myeloid composition and/or differentiation state, rather than intrinsic transcriptional upregulation of individual inflammatory mediators, underlies the observed immune phenotype in *RANBP2* mutation carriers. At baseline, circulating PBMCs did not display transcriptomic features of chronic inflammation or a basal interferonopathy, as key inflammatory genes, including TNF-α, and representative interferon-stimulated genes were not elevated at the individual transcript level (**appendix Figure S3C**). Instead, cell-type deconvolution of the bulk RNA-sequencing data using CIBERSORTx and the LM22 leukocyte reference matrix^18^ identified a relative enrichment of an M1-like macrophage signature in samples from *RANBP2* mutation carriers treated with R848, supporting a compositional or differentiation-state shift (**Figure 2D**).

### Enrichment of CXCR3-high circulating myeloid cells in ANE1

Circulating myeloid cells were identified by expression of CD14 and further resolved into subsets based on chemokine receptor expression (CCR1, CCR2, CCR5, CXCR3, and CX3CR1), reflecting differences in migratory behavior and inflammatory responsiveness (**Figure 3A, appendix Figure S4A**). Classical monocytes were defined by high CD14 expression with CCR1 and CCR2 positivity (clusters 6-8), whereas nonclassical monocytes were inferred based on lower CD14 expression and higher CX3CR1 expression, consistent with known phenotypic features of this subset^19^. CCR5 and CXCR3 expression was evaluated to identify myeloid populations with heightened inflammatory chemokine responsiveness, and CD56 was included as a marker of activated inflammatory myeloid subsets^20–22^. The overall frequency of CD14⁺ myeloid cells did not differ significantly between *RANBP2* mutation carriers and noncarrier controls, indicating preservation of the global myeloid compartment (**appendix Figure S4B**).

**Figure 3.**
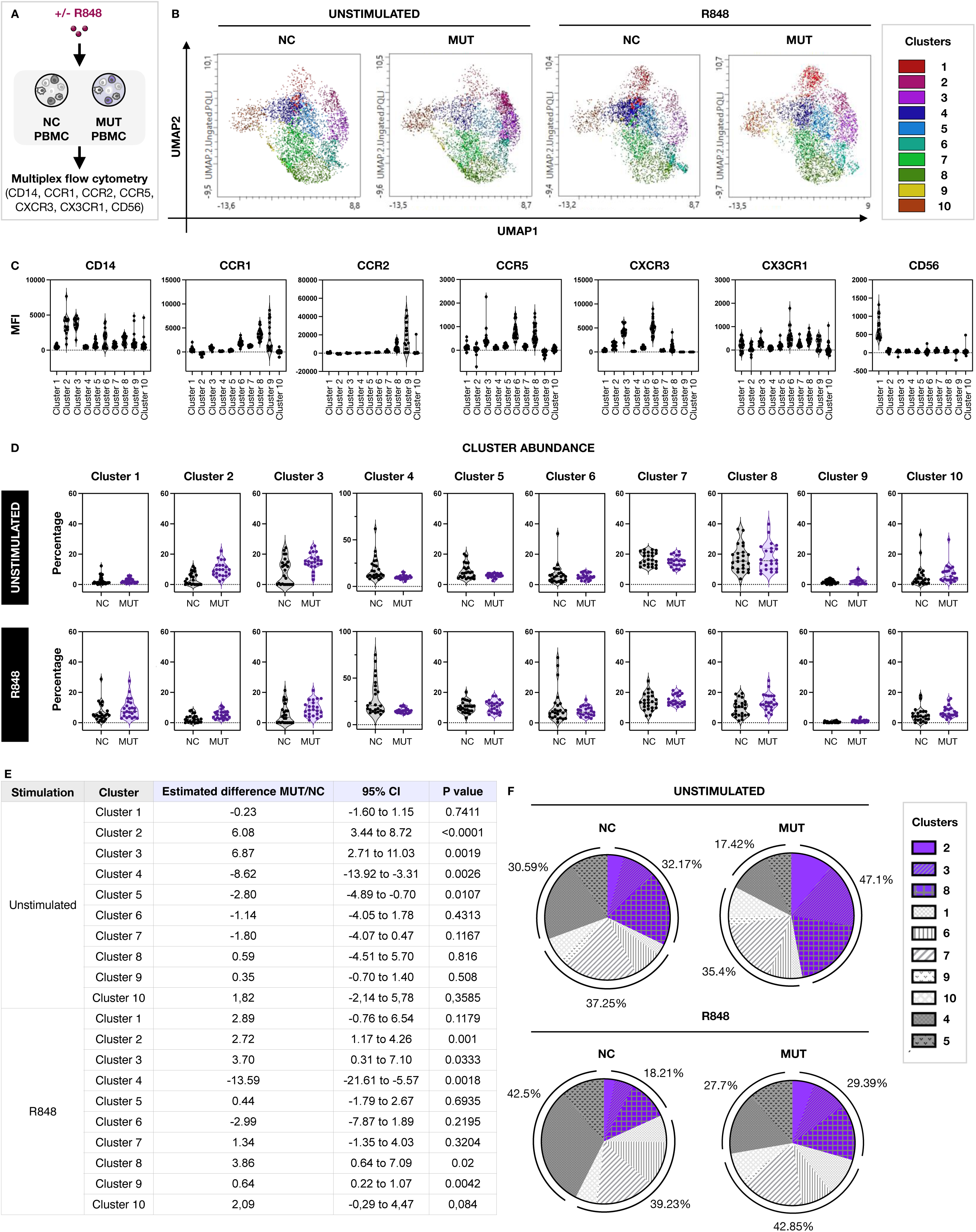
**Unsupervised multiparameter flow cytometry analysis of CD14+ myeloid cells according to chemokine receptor expression.** (A) Experimental design. PBMCs from RANBP2 c.1754C>T (p.Thr585Met) mutation carriers (MUT) and noncarriers (NC), under either unstimulated conditions or stimulated with R848, were analysed by multiparameter flow cytometry analysis of CD14, CCR1, CCR2, CCR5, CXCR3, CX3CR1, and CD56 expression. The gating strategy is provided in **appendix Figure S4C**. (B) Uniform Manifold Approximation and Projection (UMAP) visualization of CD14⁺ myeloid cells, identifying 10 phenotypically distinct clusters across NC and MUT samples under unstimulated and R848-stimulated conditions. UMAP clustering was performed on concatenated, normalised, and batch-corrected data and was used for visualisation only. (C) Marker expression profiles for each cluster in unstimulated WT samples, shown as mean fluorescence intensity (MFI). Each point represents an individual participant. (D) Violin plots display the distribution of per-patient cluster frequencies with medians (dashed line) and interquartile ranges (dotted lines). (E) Statistical analysis. Between-group comparisons of per-participant cluster frequencies were performed using unpaired Welch’s t test (GraphPad Prism, version 10). Estimated mean differences with 95% confidence intervals and exact P values are shown; similar results were obtained using a nonparametric Mann-Whitney sensitivity analysis. (F) Pie charts summarize the relative distribution of CD14⁺ myeloid clusters in MUT and NC groups.

Unsupervised clustering of CD14⁺ cells identified 10 phenotypically distinct populations that were quantified across all conditions (**Figure 3B-C, appendix Figure S4C-D; appendix p3**). The principal difference associated with *RANBP2* mutation status was an enrichment of three CXCR3-high CD14-high myeloid populations, which together accounted for most of the observed shift within the CD14⁺ compartment. Two CXCR3-high CD14-high clusters lacking CCR2 expression (clusters 2 and 3) were consistently expanded in mutation carriers under both unstimulated and stimulated conditions, with increases at baseline of cluster 2 (CXCR3-high CD14-high) accounting for the predominant change (estimated difference, 6.08 percentage points; 95% CI, 3.44 to 8.72; P<0.0001). Cluster 3, which exhibited moderately higher expression of CX3CR1, CCR5, and CCR1, likely represented a more advanced stage of monocyte differentiation^15,23^. A third CXCR3-high CD14-high population (cluster 8), that additionally expressed high levels of CCR1 and CCR2, was selectively enriched following innate immune stimulation with R848 (**Figure 3D, E**).

The expansion of CXCR3-high subsets was accompanied by a reciprocal reduction in clusters characterized by lower chemokine receptor expression, which predominated in unstimulated control samples (clusters 4 and 5). All remaining clusters showed minimal or no differences between groups, including rare CD56⁺ or CCR5-high CD14⁺ populations (**Figure 3F**).

Taken together, these data support a model in which *RANBP2* mutation status is associated with enrichment of CXCR3-high CD14-high monocytes at baseline, and CXCR3-high CD14-high monocytes with elevated inflammatory markers after innate immune stimulation (cluster 8).

### Association of CXCR3-high myeloid cells and TNF-α production with disease burden in ANE1

Among the 23 *RANBP2* mutation carriers, disease burden was assessed retrospectively using a composite score integrating disease recurrence (number of ANE episodes), cognitive impairment, clinician-rated functional disability (mRS), and long-term support requirements, with higher scores indicating greater burden (**appendix Table S1**). Because clinical data were collected retrospectively and severity was assessed using functional outcomes rather than standardized acute radiologic criteria, this measure reflects long-term functional outcome rather than initial disease severity and was evaluated at variable intervals following the most recent episode (median, 9 years). For analysis, participants were grouped into ordered severity categories, which were treated as an ordinal variable.

The frequency of a CXCR3-high CD14-high myeloid population (cluster 2) showed a strong positive association with increasing disease burden under both unstimulated and innate immune-stimulated conditions. Similar, associations were observed for a second CXCR3-high CD14-high population (cluster 3), whereas no significant association with disease burden was detected for a third CXCR3-high subset (cluster 8) (**Figure 4A-B**).

**Figure 4.**
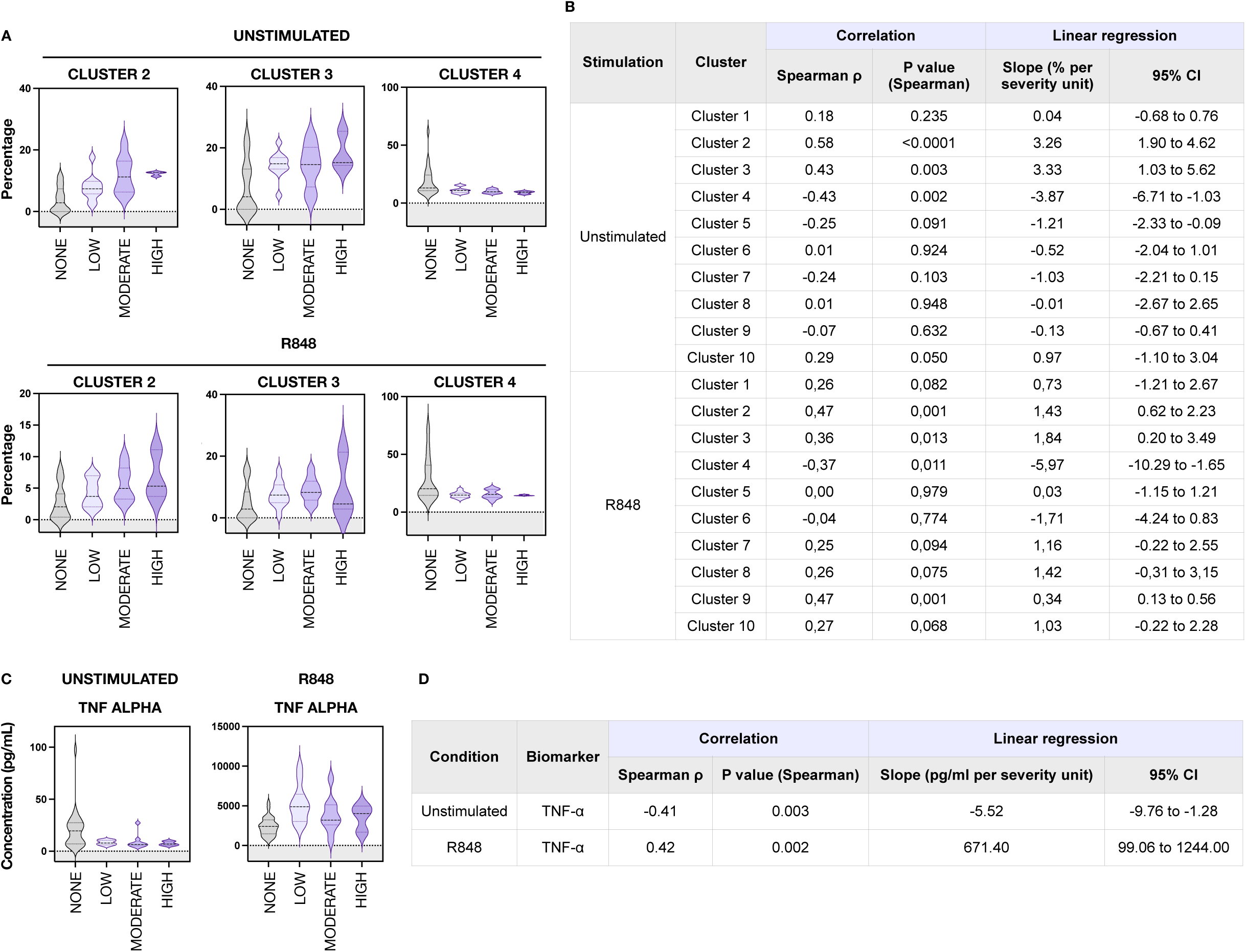
Association of myeloid cluster abundance and TNF-α production with clinical severity. (A) Violin plots show the distribution of clusters 2, 3, and 4 frequencies according to the clinical severity score, under unstimulated conditions and after stimulation with resiquimod (R848). Each point represents an individual participant. For visualization, ordinal severity scores (0-3) are displayed as categorical labels (None, Low, Moderate, High). (B) Statistical analysis of the association between cluster frequencies and clinical severity using Spearman rank correlation and linear regression models; effect estimates, 95% confidence intervals, and exact P values for panel A. (C) Violin plots show the distribution of TNF-α concentrations in unstimulated and R848-stimulated samples according to the clinical severity score. (D) Statistical analysis of the association between TNF-α production and clinical severity for panel C. Violin plots display medians (dashed lines) and interquartile ranges (dotted lines). Statistical analyses were performed using GraphPad Prism (version 10).

In parallel, PBMC TNF-α production varied with long-term clinical burden, with progressively reduced basal secretion and increased TNF-α induction in response to innate immune stimulation with increasing severity (**Figure 4C-D**).

Together, these findings indicate that increasing disease burden in ANE1 is associated with enrichment of CXCR3-high circulating myeloid states and dysregulated inflammatory responsiveness.

### Subnuclear reorganization of RANBP2 and histone deacetylases in ANE1

Myeloid cells arise from a common hematopoietic precursor and undergo sequential differentiation stages regulated by coordinated transcriptional and epigenetic programs ^16,23,24^. Nuclear pore complexes participate in these processes by controlling the spatial organization and post-translational modification of transcriptional regulators. RANBP2 has previously been shown to be required for the nuclear transport and biochemical modification of histone deacetylases HDAC3 and HDAC4^25,26^, prompting us to examine the subnuclear distribution of these proteins in participants.

Immunofluorescence labeling of HDAC3 and HDAC4 was performed in monocyte-derived macrophages from one *RANBP2* mutation carrier and one matched control (**appendix Figure S5A-D**). In control cells, HDAC3 and HDAC4 showed a predominantly nuclear and diffuse staining pattern. In contrast, cells from the mutation carrier exhibited redistribution of both proteins into discrete nuclear foci, which appeared to localize preferentially within euchromatic regions, as assessed by nuclear morphology and DNA staining. Such redistribution may reflect aberrant targeting or retention of these chromatin regulators within transcriptionally active nuclear compartments and suggests a potential link between *RANBP2* mutations and altered transcriptional control during myeloid differentiation.

## Discussion

Our study reveals that familial ANE is characterized by a dysregulated inflammatory state involving a reduced basal production of inflammatory mediators coupled to exaggerated TNF-α production following innate immune stimulation. Additionally, we uncover that this immune signature tracks with long-term disease burden, further defined by expansion of CXCR3-high CD14-high myeloid cells that likely mediate these dysregulated TNF-α responses.

These findings have important clinical translation. Current clinical evidence suggests that ANE results from a hyperacute cytokine storm, usually triggered by a viral infection, resulting in blood-brain barrier disruption and metabolic dysfunction within the central nervous system^27,28^. Alongside IL-6, elevated levels of TNF-α in the serum and/or cerebrospinal fluid are reported in patients with acute ANE^29,30^. Blockade of the IL-6 has shown promise in case reports and small series, particularly if administered early^31^. The exaggerated TNF-α response observed in our study raises the possibility that similar targeted cytokine modulation could be explored. However, given the complex role of TNF-α in host defense, such approaches would require careful evaluation. Importantly, the absence of clinical evidence of ongoing inflammation between episodes, together with the apparent hyporesponsiveness at baseline, should caution against the role of TNF-α blockade as relapse prevention/disease modifying treatment, the next major challenge in managing ANE1.

Our findings further demonstrate that carriers of *RANBP2* mutations exhibit global reprogramming of the myeloid compartment, characterized by blunted baseline responsiveness and exaggerated TNF-α production in response to TLR7/8 stimulation. This response profile recapitulates features of antiviral innate immune activation and provides a mechanistic clue for the longstanding clinical observation that ANE episodes are frequently precipitated by viral infection^32^. Additional mechanistic studies will be needed to define how *RANBP2* mutations lead to transcriptional reprogramming in myeloid cells. Whether similar pathways operate in sporadic, non-genetic forms of ANE also remains to be determined. Importantly, these pathways are currently being targeted to good clinical effects with HDAC inhibitors, such as Givinostat in Duchene muscular dystrophy^33^.

CXCR3 is a chemokine receptor for CXCL10, a key downstream effector of type I and type II interferon signaling. Expression of CXCR3 on myeloid cells has been linked to enhanced trafficking to inflamed tissues and increased tissue infiltration including transmigration^34^. Additionally, the observed CX3CR1 expression profiles are consistent with an enrichment of both early and intermediate monocyte subsets with tissue-homing capacity (clusters 2 and 3)^16,19^. However, more detailed phenotypic characterization, including incorporation of canonical monocyte markers such as CD16, will be required to properly identify the myeloid populations expanded in ANE1 and to assess their functional and migratory properties. Although immune cell infiltration of the central nervous system (CNS) has been described in other inflammatory neurologic conditions, direct evidence for BBB transmigration of circulating myeloid cells in ANE1 is currently lacking. Further studies will therefore be needed to determine whether CXCR3high myeloid cells display increased CNS tropism and whether they represent a principal source of TNF-α in vivo.

This study has several limitations. First, clinical data were collected retrospectively, with disease burden assessed by clinician-rated functional impairment rather than standardized acute radiologic criteria. As a consequence, severity in this study reflects long-term functional outcome rather than initial disease burden, and was assessed at variable intervals after the most recent episode, thereby limiting normalization across participants.

Second, the cohort design required participant travel, which limited the inclusion of individuals with the most severe disease. Among participants with one or more episodes who were enrolled, 3 of 12 (25%) experienced recurrence. When all invited persons were considered, the recurrence rate was higher (8 of 18, 44%), indicating underrepresentation of individuals with severe recurrent disease.

Finally, what remains unresolved is the incomplete penetrance of *RANBP2* mutations, which has been estimated at approximately 40%^10^. In this cohort, 12 of 23 mutation carriers (52%) reported at least one ANE episode. Our data also do not explain why ANE occurs predominantly in childhood (median age of onset 5-10 years). Together, these observations indicate that *RANBP2* mutations alone are insufficient to determine ANE manifestation or recurrence. Identification of additional genetic or environmental modifiers will depend on further studies and require larger international collaborative cohorts.

## Author contributions

Concept and design (Lim; Picot; Meyer; Arhel), Acquisition of data (Desgraupes; Boireau; Khalil; Nisole; Bolloré; Meyer; Arhel), Analysis and interpretation of data (Desgraupes; Boireau; Aouinti; Lim; Picot; Meyer; Arhel), Statistical analysis and bioinformatics (Desgraupes; Aouinti; Picot; Arhel), Patient recruitment and clinical coordination (Khalil; Picot; Meyer), Laboratory assays and resources (Nisole; Bolloré; Tuaillon), Clinical leadership and expert consultation (Barbaria; Barzaghi; Dilena; Boon; Lunsing; Westerholm-Ormio; Deiva; Bakker; Kuijpers; Yeh; Lim; Picot; Meyer), Funding acquisition (Meyer; Arhel), Drafting and revision of the manuscript (all authors)

## Data Availability

All data produced in the present study are available upon reasonable request to the authors.

## Acknowledgements

The authors wish to thank the study participants and all ANE families for their generous contribution to research. The study was conducted with funds from the Agence Nationale de la Recherche (ANR-23-CE15-0005-01) and ANRS-MIE (ECTZ209411) to N.J.A, a CHU Montpellier Tremplin grant to P.M. and N.J.A. Family travel was financed by the Grayson Legacy Support Fund. RNA sequencing and computational analysis was performed by the McGill Genome Center (Canada). Flow cytometry and microscopy was carried out at Montpellier Resources Imagerie (MRI). We extend special thanks to Kim Smith from ANE International for assistance in organising family travel to Montpellier. We also thank the promotor for the study (CHU Montpellier) and Sylvie Brochot, Quentin Plassais, Charlotte Kaan, Hélène Lenden Hasse and Amine Zaamou from the Research Innovation Department. We also acknowledge Joana Da Silva Pissarra, medical writer. We are grateful to all participating medical personnel for their assistance with patient recruitment and blood sampling.

## References

1. Fazal A, Reinhart K, Huang S, et al. Reports of Encephalopathy Among Children with Influenza-Associated Mortality — United States, 2010–11 Through 2024–25 Influenza Seasons. MMWR Morb Mortal Wkly Rep. 2025;74(6):91–95. doi:10.15585/mmwr.mm7406a3

2. Uyeki TM. Pediatric Influenza-Associated Acute Necrotizing Encephalopathy—Gaps Need to Be Addressed. JAMA. 2025;334(8):677. doi:10.1001/jama.2025.13003

3. Silverman A, Walsh R, Santoro JD, et al. Influenza-Associated Acute Necrotizing Encephalopathy in US Children. JAMA. 2025;334(8):692. doi:10.1001/jama.2025.11534

4. Sakuma H, Thomas T, Debinski C, et al. International consensus definitions for infection-triggered encephalopathy syndromes. Dev Med Child Neurol. 2025;67(2):195–207. doi:10.1111/dmcn.16067

5. Chatur N, Yea C, Ertl-Wagner B, Yeh EA. Outcomes in influenza and *RANBP2* mutation-associated acute necrotizing encephalopathy of childhood. Develop Med Child Neuro. 2022;64(8):1008–1016. doi:10.1111/dmcn.15165

6. Levine JM, Ahsan N, Ho E, Santoro JD. Genetic Acute Necrotizing Encephalopathy Associated with RANBP2: Clinical and Therapeutic Implications in Pediatrics. Multiple Sclerosis and Related Disorders. 2020;43:102194. doi:10.1016/j.msard.2020.102194

7. Shukla P, Mandalla A, Elrick MJ, Venkatesan A. Clinical Manifestations and Pathogenesis of Acute Necrotizing Encephalopathy: The Interface Between Systemic Infection and Neurologic Injury. Front Neurol. 2022;12:628811. doi:10.3389/fneur.2021.628811

8. Okumura A, Mizuguchi M, Kidokoro H, et al. Outcome of acute necrotizing encephalopathy in relation to treatment with corticosteroids and gammaglobulin. Brain and Development. 2009;31(3):221–227. doi:10.1016/j.braindev.2008.03.005

9. Koh JC, Murugasu A, Krishnappa J, Thomas T. Favorable Outcomes With Early Interleukin 6 Receptor Blockade in Severe Acute Necrotizing Encephalopathy of Childhood. Pediatric Neurology. 2019;98:80–84. doi:10.1016/j.pediatrneurol.2019.04.009

10. Neilson DE, Adams MD, Orr CMD, et al. Infection-Triggered Familial or Recurrent Cases of Acute Necrotizing Encephalopathy Caused by Mutations in a Component of the Nuclear Pore, RANBP2. The American Journal of Human Genetics. 2009;84(1):44–51. doi:10.1016/j.ajhg.2008.12.009

11. Desgraupes S, Decorsière A, Perrin S, et al. The genetic driver of Acute Necrotizing Encephalopathy, RANBP2, regulates the inflammatory response to Influenza A virus infection. Nat Commun. Published online February 6, 2026. doi:10.1038/s41467-026-69288-1

12. Leplat M, Desgraupes S, Beucher G, et al. ANEmone: The 1st Science & Families conference on acute necrotizing encephalopathy raises hopes for an ultra-rare genetic disease. Virologie. 2025;29(6):431–434. doi:10.1684/vir.2025.1117

13. Gouy B, Decorsière A, Desgraupes S, et al. Rapid and inexpensive bedside diagnosis of RAN binding protein 2-associated acute necrotizing encephalopathy. Front Neurol. 2023;14:1282059. doi:10.3389/fneur.2023.1282059

14. Westendorp WF, Bakker DP, Duijkers FAM, Kuijpers T. Clinical Reasoning: A 2-Year-Old Girl With Acute Encephalopathy After Febrile Systemic Illness. Neurology. 2025;105(5):e213970. doi:10.1212/WNL.0000000000213970

15. Wacleche VS, Cattin A, Goulet JP, et al. CD16+ monocytes give rise to CD103+RALDH2+TCF4+ dendritic cells with unique transcriptional and immunological features. Blood Advances. 2018;2(21):2862–2878. doi:10.1182/bloodadvances.2018020123

16. Ancuta P, Liu KY, Misra V, et al. Transcriptional profiling reveals developmental relationship and distinct biological functions of CD16+ and CD16- monocyte subsets. BMC Genomics. 2009;10(1):403. doi:10.1186/1471-2164-10-403

17. Frankenberger M, Hofer TPJ, Marei A, et al. Transcript profiling of CD 16-positive monocytes reveals a unique molecular fingerprint. Eur J Immunol. 2012;42(4):957–974. doi:10.1002/eji.201141907

18. Steen CB, Liu CL, Alizadeh AA, Newman AM. Profiling Cell Type Abundance and Expression in Bulk Tissues with CIBERSORTx. In: Kidder BL, ed. Stem Cell Transcriptional Networks. Vol 2117. Methods in Molecular Biology. Springer US; 2020:135–157. doi:10.1007/978-1-0716-0301-7_7

19. Auffray C, Fogg D, Garfa M, et al. Monitoring of Blood Vessels and Tissues by a Population of Monocytes with Patrolling Behavior. Science. 2007;317(5838):666–670. doi:10.1126/science.1142883

20. Grip O, Bredberg A, Lindgren S, Henriksson G. Increased subpopulations of CD16+ and CD56+ blood monocytes in patients with active Crohn’s disease: Inflammatory Bowel Diseases. 2007;13(5):566–572. doi:10.1002/ibd.20025

21. Sconocchia G, Keyvanfar K, El Ouriaghli F, et al. Phenotype and function of a CD56+ peripheral blood monocyte. Leukemia. 2005;19(1):69–76. doi:10.1038/sj.leu.2403550

22. Krasselt M, Baerwald C, Wagner U, Rossol M. CD56+ monocytes have a dysregulated cytokine response to lipopolysaccharide and accumulate in rheumatoid arthritis and immunosenescence. Arthritis Res Ther. 2013;15(5):R139. doi:10.1186/ar4321

23. Sunderkötter C, Nikolic T, Dillon MJ, et al. Subpopulations of Mouse Blood Monocytes Differ in Maturation Stage and Inflammatory Response. The Journal of Immunology. 2004;172(7):4410–4417. doi:10.4049/jimmunol.172.7.4410

24. Randolph GJ, Sanchez-Schmitz G, Liebman RM, Schäkel K. The CD16(+) (FcgammaRIII(+)) subset of human monocytes preferentially becomes migratory dendritic cells in a model tissue setting. J Exp Med. 2002;196(4):517–527. doi:10.1084/jem.20011608

25. Guglielmi V, Lam D, D’Angelo MA. Nucleoporin Nup358 drives the differentiation of myeloid-biased multipotent progenitors by modulating HDAC3 nuclear translocation. Science Advances. 2024;10(23):eadn8963. doi:10.1126/sciadv.adn8963

26. Kirsh O, Seeler JS, Pichler A, et al. The SUMO E3 ligase RanBP2 promotes modification of the HDAC4 deacetylase. EMBO J. 2002;21(11):2682–2691. doi:10.1093/emboj/21.11.2682

27. Dale RC, Thomas T, Patel S, et al. CSF neopterin and quinolinic acid are biomarkers of neuroinflammation and neurotoxicity in FIRES and other infection-triggered encephalopathy syndromes. Ann Clin Transl Neurol. 2023;10(8):1417–1432. doi:10.1002/acn3.51832

28. Mizuguchi M, Shibata A, Kasai M, Hoshino A. Genetic and environmental risk factors of acute infection-triggered encephalopathy. Front Neurosci. 2023;17:1119708. doi:10.3389/fnins.2023.1119708

29. Ichiyama T, Endo S, Kaneko M, Isumi H, Matsubara T, Furukawa S. Serum cytokine concentrations of influenza-associated acute necrotizing encephalopathy. Pediatrics International. 2003;45(6):734–736. doi:10.1111/j.1442-200X.2003.01822.x

30. Ichiyama T, Isumi H, Ozawa H, Matsubara T, Morishima T, Furukawa S. Cerebrospinal Fluid and Serum Levels of Cytokines and Soluble Tumor Necrosis Factor Receptor in Influenza Virus-associated Encephalopathy. Scandinavian Journal of Infectious Diseases. 2003;35(1):59–61. doi:10.1080/0036554021000026986

31. Fischell SZ, Fischell J, Kliot T, Tumulty J, Thompson SJ, Raees MQ. Case report: Acute necrotizing encephalopathy: a report of a favorable outcome and systematic meta-analysis of outcomes with different immunosuppressive therapies. Front Neurol. 2023;14:1239746. doi:10.3389/fneur.2023.1239746

32. Mizuguchi M, Yamanouchi H, Ichiyama T, Shiomi M. Acute encephalopathy associated with influenza and other viral infections. Acta Neurol Scand. 2007;115(s186):45–56. doi:10.1111/j.1600-0404.2007.00809.x

33. Mercuri E, Vilchez JJ, Boespflug-Tanguy O, et al. Safety and efficacy of givinostat in boys with Duchenne muscular dystrophy (EPIDYS): a multicentre, randomised, double-blind, placebo-controlled, phase 3 trial. Lancet Neurol. 2024;23(4):393–403. doi:10.1016/S1474-4422(24)00036-X

34. Niu F, Liao K, Hu G, Moidunny S, Roy S, Buch S. HIV Tat-Mediated Induction of Monocyte Transmigration Across the Blood–Brain Barrier: Role of Chemokine Receptor CXCR3. Front Cell Dev Biol. 2021;9:724970. doi:10.3389/fcell.2021.724970

