## Appendix for "An Inflammatory Signature Associated with Genetic Predisposition to Acute Necrotizing Encephalopathy"

**Methods……………………** ……………………………………………………………………p2

**Figures……………………**…………………………………………………………………...p5

Figure S1………………………………………………………………………………….p5

Figure S2………………………………………………………………………………….p5

Figure S3………………………………………………………………………………….p6

Figure S4………………………………………………………………………………….p7

Figure S5………………………………………………………………………………….p8

**Tables……………………**…………………………………………………………………p9

Table S1…………………………………………………………………………………..p9

#### Methods

##### Allelic RTqPCR

RNA was extracted from peripheral-blood lymphocytes and subjected to RANBP2-specific reverse transcription, as previously described[19]. Wild-type and c.1754C>T *RANBP2* alleles were quantified by allele-specific quantitative PCR using discriminating primers, together with primers detecting total *RANBP2*. Assay performance was validated using HEK-293T cells expressing wild-type, mutant, or both *RANBP2* alleles.

##### Inflammatory marker quantification

The primary outcome was the expression of pro-inflammatory markers, assessed under basal conditions and following *ex vivo* stimulation. Cytokine and chemokine concentrations in PBMC culture supernatants were quantified using a magnetic bead–based multiplex assay (20-plex Human Inflammation Panel, PROCARTAPLEX®, Thermo Fisher Scientific) and analyzed on a Luminex MAGPIX® instrument (xMAP technology), according to the manufacturer's instructions. The limits of detection ranged from <1 to 5 000 000 pg/mL, depending on the analyte. Standard curves were generated for each cytokine to calculate concentrations, and a minimum of 50 beads per analyte were acquired for analysis. Concentrations exceeding the upper limit of quantification were assigned an arbitrary value corresponding to the highest point of the standard curve for the respective biomarker. All multiplex cytokine measurements using the Luminex platform were performed centrally at the Clinical Epidemiology and Biostatistics Unit (URCE, SIMED, Montpellier University Hospital). IL-6 concentrations frequently exceeded the upper detection limit and were therefore confirmed using ELISA (Proteintech KE00385).

##### RNAseq

Analysis was performed on *RANBP2* mutation carriers and related noncarriers only. Adaptor sequences and low quality score bases from the 3′ end (Phred score < 30) were first trimmed using *Trimmomatic*[13]. Any read shorter than 32 bp is further discarded. The resulting reads were aligned to the GRCh38 assembly of the *Homo sapiens* reference genome, using *STAR*[14]. Read counts were obtained using *HTSeq*[15]with parameters *-m intersection-nonempty -stranded=reverse*. For all downstream analyses, we excluded lowly-expressed genes with an average read count lower than 10 across all samples. Raw counts were normalized using *edgeR*'s TMM algorithm[16] and were then transformed to log2-counts per million (log2CPM) using the *voom* function implemented in the *limma* R package[17]. Differential gene expression was assessed using a linear model via *lmFit* (method = "robust"), including Sex, Age, and Batch as covariates. Nominal p-values were corrected for multiple testing using the Benjamini-Hochberg method. Genes with an adjusted p-value < 0.05 were considered differentially expressed. Gene set enrichment analysis based on pre-ranked gene list by t-statistic was performed using the R package *fgsea* (http://bioconductor.org/packages/fgsea/).

RNA sequencing data were analyzed independently using standard differential expression pipelines (edgeR and DESeq2), with comparisons performed between statistically independent sample groups.

**Flow-Cytometry**

Samples were acquired on a BD LSR Fortessa flow cytometer using standardized acquisition and compensation procedures across experiments. Flow-cytometry data were analyzed using unsupervised dimensionality-reduction and clustering approaches implemented in FlowJo (version 10). Cluster frequencies and marker expression patterns were quantified for each participant and compared across experimental conditions. Statistical analyses were performed using GraphPad Prism (version 10).

**Cellular imaging**

Monocytes were isolated by adherence of PBMCs to plastic for 45 minutes, as previously described[18]. Adherent cells were differentiated into macrophages for 7 days in GM-CSF-supplemented medium. Cells were fixed, permeabilized, and incubated with primary rabbit anti-HDAC3 or -HDAC4 antibodies (Santa cruz, sc-11417, sc-11418, 1/200), followed by secondary antibodies (Thermofisher Scientific, A-21429, 1/1000) and Hoechst. Images were acquired by LSM800 confocal microscopy and analyzed with Fiji software.

**Statistical analyses for secondary analyses**

Statistical analyses were performed using R software (version 4.5.0) and Graphpad Prism (version 10).

For secondary analyses of flow cytometry and imaging data, between-group comparisons of per-participant cluster frequencies were performed using unpaired Welch's t test in GraphPad Prism, version 10. Estimated mean differences with 95% confidence intervals and exact P values are reported. Sensitivity analyses using the Mann–Whitney test yielded similar results. Associations between cluster frequencies and disease burden were evaluated using Spearman rank correlation and linear regression models; effect estimates, 95% confidence intervals, and exact P values are provided.

For RNA-sequencing analyses, differential gene expression was assessed using linear modeling in R with the limma package (lmFit, robust option), including sex, age, and batch as covariates. Nominal P values were adjusted for multiple testing using the Benjamini–Hochberg method. Genes with an adjusted P value less than 0.05 were considered differentially expressed. Gene-set enrichment analysis was performed on a preranked gene list (by t statistic) using the fgsea package.

#### Figure S1

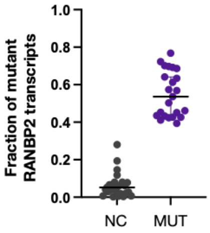


**Figure S1. Allele-specific expression of *RANBP2*.**

Mutant RANBP2 allelic expression in PBMCs under basal conditions, quantified by targeted RT-PCR with allele-discriminant quantitative PCR and expressed as the mutant allelic fraction (Q_MUT / [Q_NC + Q_MUT], from $2^{-Ct}$ values). Each dot denotes one biological sample (n = 25 NC; n = 23 ANE1); bars show the median and 95% confidence intervals. Samples failing quality-control criteria were excluded.

#### Figure S2

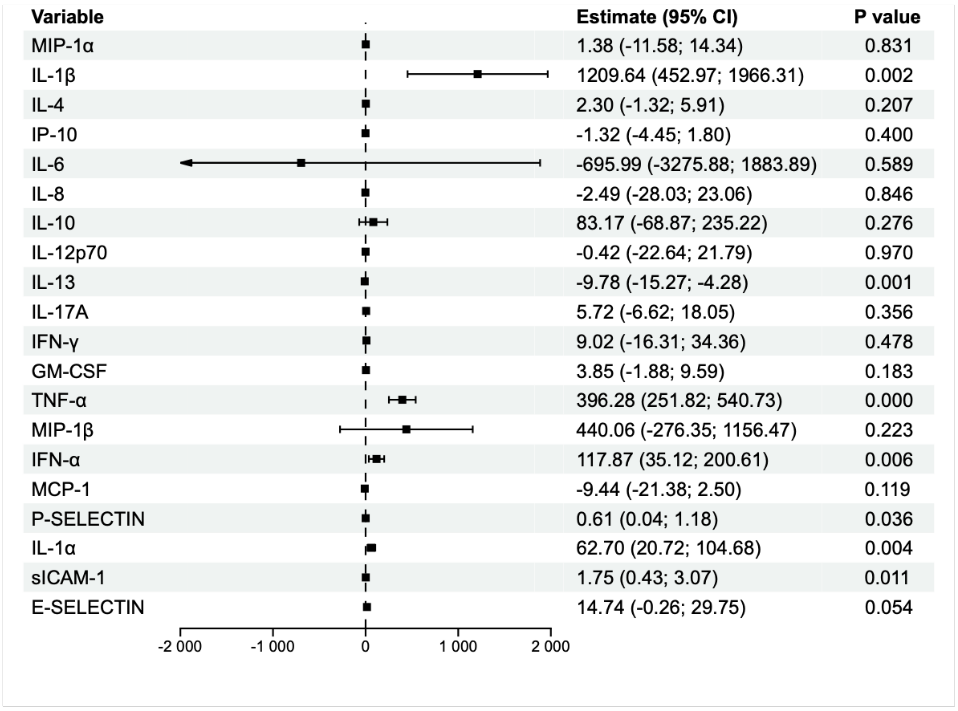


| Variable | Estimate (95% CI) | P value |
|---|---|---|
| MIP-1α | 1.38 (-11.58; 14.34) | 0.831 |
| IL-1β | 1209.64 (452.97; 1966.31) | 0.002 |
| IL-4 | 2.30 (-1.32; 5.91) | 0.207 |
| IP-10 | -1.32 (-4.45; 1.80) | 0.400 |
| IL-6 | -695.99 (-3275.88; 1883.89) | 0.589 |
| IL-8 | -2.49 (-28.03; 23.06) | 0.846 |
| IL-10 | 83.17 (-68.87; 235.22) | 0.276 |
| IL-12p70 | -0.42 (-22.64; 21.79) | 0.970 |
| IL-13 | -9.78 (-15.27; -4.28) | 0.001 |
| IL-17A | 5.72 (-6.62; 18.05) | 0.356 |
| IFN-γ | 9.02 (-16.31; 34.36) | 0.478 |
| GM-CSF | 3.85 (-1.88; 9.59) | 0.183 |
| TNF-α | 396.28 (251.82; 540.73) | 0.000 |
| MIP-1β | 440.06 (-276.35; 1156.47) | 0.223 |
| IFN-α | 117.87 (35.12; 200.61) | 0.006 |
| MCP-1 | -9.44 (-21.38; 2.50) | 0.119 |
| P-SELECTIN | 0.61 (0.04; 1.18) | 0.036 |
| IL-1α | 62.70 (20.72; 104.68) | 0.004 |
| sICAM-1 | 1.75 (0.43; 3.07) | 0.011 |
| E-SELECTIN | 14.74 (-0.26; 29.75) | 0.054 |

**Figure S2. Fold increase in cytokine secretion following R848 stimulation.**

Fold changes in cytokine secretion (R848-stimulated divided by unstimulated) for 20 cytokines measured in PBMCs from individuals carrying RANBP2 mutations and from age- and sex-matched noncarrier controls. Group comparisons were performed using linear mixed-effects models with group as a fixed effect and matched pairs as random effects (two-sided α = 0.05).

### Figure S3

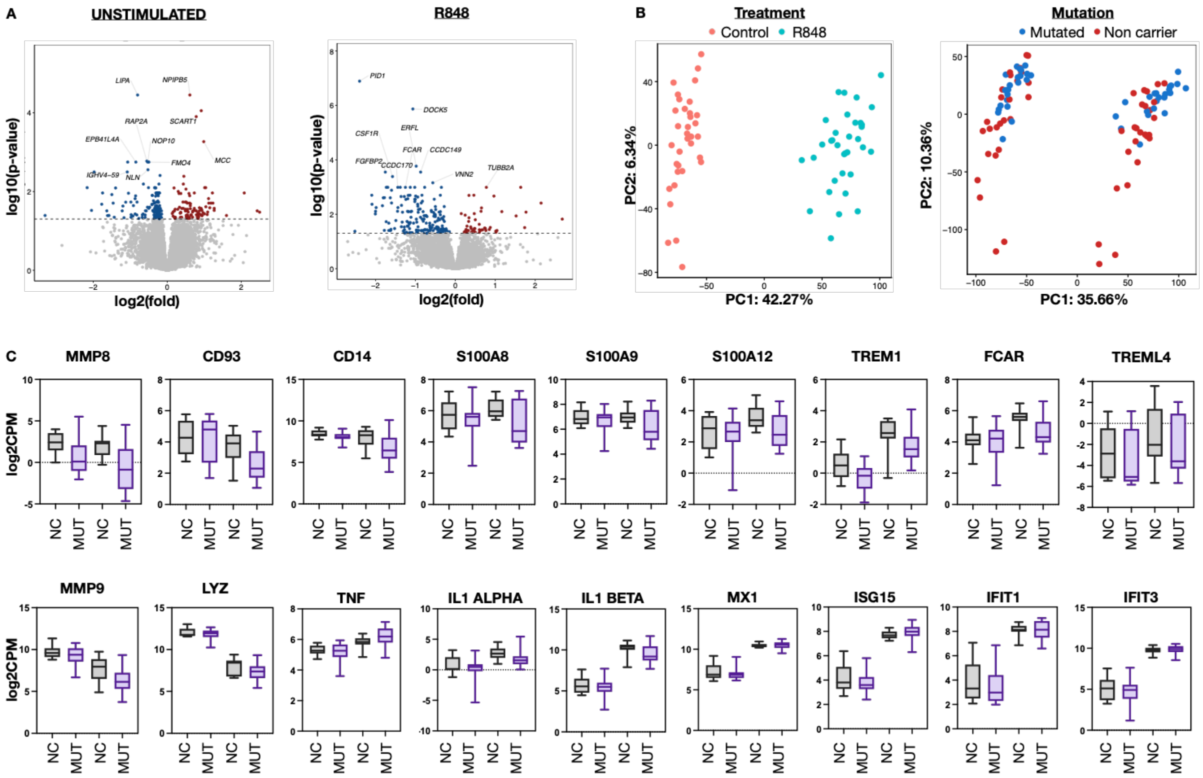


**Figure S3. Extended data for RNA-sequencing analysis.**

(A) Volcano plots of differential expression in unstimulated and R848-stimulated PBMCs from RANBP2 mutation carriers (MUT) versus noncarriers (NC).

(B) Principal-component analysis (PCA) of normalized RNA-sequencing data from PBMCs shows clear separation of samples according to stimulation condition (unstimulated vs. R848), with partial segregation by RANBP2 mutation status. Plots display the first two principal components (PC1 and PC2), summarizing global transcriptional variation across samples.

(B) Normalized expression (log2CPM) of selected genes for with a classical monocyte transcriptional signature, as well as inflammatory and interferon-response genes.

#### Figure S4

A

| Fluorophore | Marker | Provider | Reference | Clone |
|---|---|---|---|---|
| FITC | CD14 | Biolegend | 325604 | Clone HCD14 |
| APC | CD56 (NCAM) | Biolegend | 318310 | Clone HCD56 |
| A700 | CD3 | Biolegend | 344822 | Clone SK7 |
| BV421 | CXCR3 | Biolegend | 353716 | Clone G025H7 |
| BV650 | CCR5 | Biolegend | 359148 | Clone J418F1 |
| BV711 | CX3CR1 | Biolegend | 341630 | Clone 2A9-1 |
| PE-Dazzle594 | CCR2 | Biolegend | 357222 | Clone K036C2 |
| PE-Cy7 (Vio770) | CCR1 | Biolegend | 362914 | Clone 5F10B29 |

B

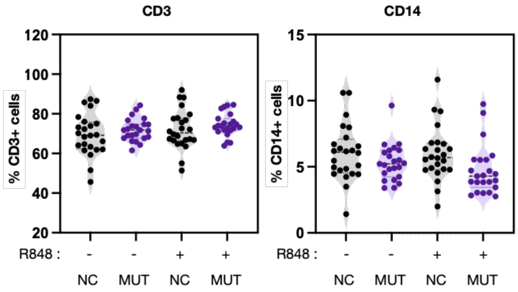


C

D

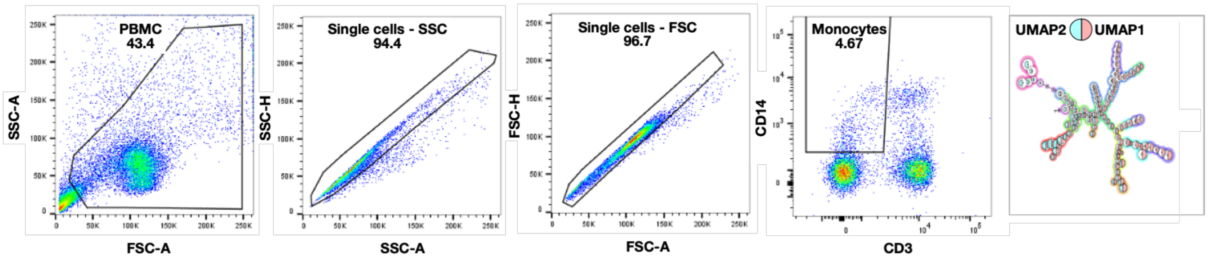


**Figure S4. Extended flow-cytometry analyses.**

(A) Antibody panel used for flow-cytometry analyses.
(B) Frequencies of $CD14^+$ monocytes and $CD3^+$ T cells across conditions; each point represents one participant.
(C) Gating strategy illustrated by representative flow-cytometry plots identifying $CD3^-CD14^+$ monocytes within PBMCs. Samples were acquired on a BD LSR Fortessa cytometer with compensation based on single-marker controls.
(D) Example of UMAP visualization with FlowSOM clustering (10 clusters) for $CD3^-CD14^+$ monocytes from R848-stimulated mutation carriers. Unsupervised analyses were performed in FlowJo v10 on equal numbers of monocytes (n=250 per group), concatenated by genotype and stimulation condition. Dimensionality reduction and clustering were applied using UMAP and FlowSOM via the Embed plugin, with cluster frequencies assessed using the Cluster Explorer plugin.

#### Figure S5

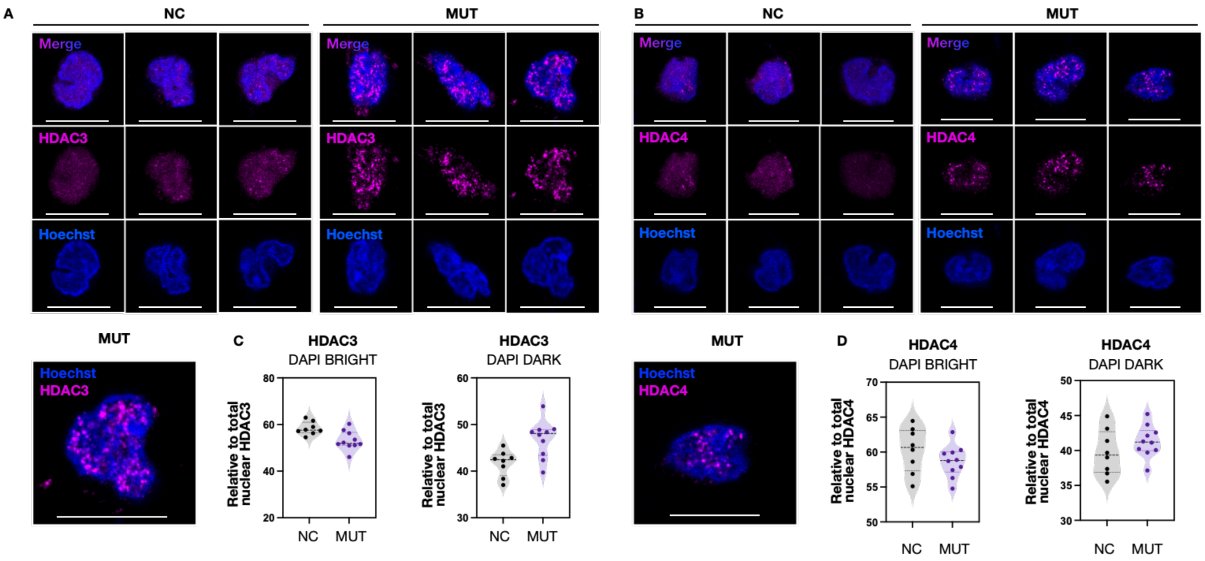


**Figure S5. Subnuclear distribution of HDAC3 and HDAC4 in monocytes**

(A-B) Representative immunofluorescence images of HDAC3 and HDAC4 (magenta) in monocytes from one RANBP2 T585M mutation carrier (MUT) and one matched control (NC). In control cells, HDAC3 and HDAC4 display predominantly diffuse nuclear localization, whereas in mutant cells both proteins form discrete nuclear foci. Scale bars, 10 µm.

(C-D) Regions exhibiting either high or low DAPI staining intensity were identified using threshold. HDAC3 and HDAC4 signals were quantified within each region. Values are expressed as a percentage of the total nuclear fluorescence, with each dot corresponding to an individual cell.

#### Table S1

| RANBP2-positive participant | No. of ANE episodes | Cognitive severity | mRS score | Special needs assessment | Composite score | Score category | Years since most recent episode |
|---|---|---|---|---|---|---|---|
| F01-P01 | 1 | 0 | 0 | 0 | 1 | Moderate | 18 |
| F01-P03 | 1 | 0 | 2 | 0 | 3 | Moderate | 2 |
| F02-P02 | 0 | 0 | 0 | 0 | 0 | Low | |
| F02-P04 | 2 | 1 | 0 | 0 | 3 | Moderate | 5 |
| F02-P05 | 1 | 0 | 0 | 0 | 1 | Moderate | 47 |
| F04-P02 | 0 | 0 | 0 | 0 | 0 | Low | |
| F05-P02 | 0 | 0 | 0 | 0 | 0 | Low | |
| F05-P03 | 3 | 1 | 2 | 1 | 7 | High | 6 |
| F06-P02 | 1 | 0 | 0 | 0 | 1 | Moderate | 7 |
| F06-P03 | 1 | 0 | 0 | 0 | 1 | Moderate | 3 |
| F06-P04 | 1 | 1 | 4 | 2 | 8 | High | 11 |
| F06-P05 | 0 | 0 | 0 | 0 | 0 | Low | |
| F07-P01 | 0 | 0 | 0 | 0 | 0 | Low | |
| F07-P03 | 1 | 0 | 0 | 0 | 1 | Moderate | 1 |
| F07-P04 | 0 | 0 | 0 | 0 | 0 | Low | |
| F09-P01 | 0 | 0 | 0 | 0 | 0 | Low | |
| F09-P02 | 1 | 1 | 0 | 1 | 3 | Moderate | 40 |
| F09-P03 | 3 | 1 | 1 | 1 | 6 | High | 19 |
| F09-P04 | 0 | 0 | 0 | 0 | 0 | Low | |
| F10-P02 | 0 | 0 | 0 | 0 | 0 | Low | |
| F10-P03 | 1 | 1 | 0 | 1 | 3 | Moderate | 11 |
| F10-P04 | 0 | 0 | 0 | 0 | 0 | Low | |
| F10-P05 | 0 | 0 | 0 | 0 | 0 | Low | |

**Table S1. ANE disease burden impact score.**

Composite score (range 0–14) summarizing ANE burden based on number of episodes, cognitive impairment severity, functional disability (mRS), and long-term support needs at inclusion, with participants classified into ordinal severity categories from no disease burden to high impact.
